# Geospatial Quantification of Antiretroviral Therapy Consumption in Tanzania (2017-2021) Using the WHO Defined Daily Dose Methodology

**DOI:** 10.64898/2025.12.06.25341745

**Authors:** Vicky Peter Manyanga, James Mwakyomo, Hillary Mushi, Meshack D. Lugoba, Veryeh Sambu, Ritah F. Mutagonda, George Musiba, Beatrice Mutayoba, Mercy Mpatwa Masuki, Prosper Njau, Werner Maokola, Raphael Z. Sangeda

**Affiliations:** Department of Medicinal Chemistry, Muhimbili University of Health and Allied Sciences, Dar es Salaam, Tanzania; Department of Pharmaceutical Microbiology, School of Pharmacy, Muhimbili University of Health and Allied Sciences, Dar es Salaam, Tanzania; Department of Pharmacognosy, School of Pharmacy, Muhimbili University of Health and Allied Sciences, Dar es Salaam, Tanzania; National AIDS and Sexually Transmitted Infections Control Programme, Ministry of Health, Dodoma, Tanzania; Department of Clinical Pharmacy and Pharmacology, School of Pharmacy, Muhimbili University of Health and Allied Sciences, Dar es Salaam, Tanzania; Department of Pharmacy, Muhimbili National Hospital, Dar es Salaam, Tanzania; National Malaria Control Program, Ministry of Health, Dodoma, Tanzania

**Keywords:** Antiretroviral therapy (ART), Defined Daily Dose (DDD), HIV, Tanzania, Drug utilisation, Dolutegravir (DTG), Pharmacoepidemiology, Geospatial analysis

## Abstract

**Objectives:** To quantify national and regional trends in antiretroviral therapy (ART) consumption in Tanzania from 2017 to 2021 using the World Health Organisation (WHO) Defined Daily Dose (DDD) methodology and to describe shifts in regimen composition during the transition to dolutegravir-based therapy.

**Methods:** A retrospective descriptive analysis was conducted using programmatic dispensing data from the National AIDS and Sexually Transmitted Infections Control Programme (NASHCoP). ART dispensing records from all reporting HIV Care and Treatment Clinics were harmonised with WHO ATC/DDD codes and linked to annual population estimates. Consumption was expressed as DDDs per 1,000 inhabitants per day. Temporal trends, therapeutic class distributions, and regional variations were analysed using Power BI and R. Short-term national forecasts until 2025 were modelled using quadratic regression.

**Results:** Between 2017 and 2021, ART consumption increased from 12.2 to 27.1 DDD/1,000/day (compound annual growth rate, 22.4%), with a five-year mean of 19.7 DDD/1,000/day. A total of 2,108,707 unique patients accounted for 39.7 million clinic visits in the study. The average number of visits per patient per year declined from 6.78 to 5.24, consistent with the expansion of the multi-month dispensing program (MM). Nucleoside/nucleotide reverse transcriptase inhibitors (NRTIs) remained the core therapeutic class, whereas the use of integrase strand transfer inhibitors (INSTIs) has increased by more than tenfold since 2019. Dolutegravir (DTG) - based combinations (TDF/3TC/DTG) have become the predominant regimen, accounting for approximately 70% of dispensed DDDs by 2021. Regional variation narrowed over time, although the highest consumption persisted in Dar es Salaam, Mwanza, and Dodoma. Forecast modelling indicated continued moderate growth through 2025.

**Conclusions:** The national ART consumption in Tanzania more than doubled between 2017 and 2021, reflecting the expansion of treatment coverage, evolution of differentiated service delivery, and the transition to DTG-based regimens. The routine computation of DDD per 1,000 inhabitants per day provides a standardised and feasible metric for monitoring ART utilisation within national HIV programs.

## 1. Introduction

The global scale-up of antiretroviral therapy (ART) over the past two decades has transformed HIV infection from a fatal disease to a manageable chronic condition in millions of people living with HIV (PLHIV). Guided by the “Treat All” policy and the UNAIDS 95–95–95 targets, countries have expanded access to ART, simplified regimens, and strengthened their treatment monitoring systems [1]. As national programs continue to mature, there is an increasing emphasis on adopting metrics that assess not only treatment coverage and clinical outcomes but also the volume, composition, and evolution of antiretroviral medicine use at the population level. Traditional indicators, such as the number of people on ART or the proportion of those who are virally suppressed, remain critical but do not fully capture changes in regimen distribution or medicine utilisation patterns within health systems [2].

The World Health Organisation (WHO) recommends expressing national antimicrobial consumption as Defined Daily Doses (DDD) per 1,000 inhabitants per day. This standardised measure enables international comparisons and the detection of unusual utilisation patterns [3]. The DDD-based approach has been widely applied to antibiotics, antivirals, and antifungals to quantify their use and support the development of rational medicine-use policies. For example, national and global studies of antimicrobial consumption have demonstrated how DDD-based metrics reveal trends in utilisation that are otherwise obscured in traditional patient-count data [4–6]. In Tanzania, a similar methodology has been used to assess the consumption of systemic antivirals and antifungals, demonstrating the feasibility of DDD-based surveillance using programmatic data [7].

As Tanzania transitioned to DTG-based first-line therapy, early programmatic experience highlighted favourable adherence and acceptability among people living with HIV, assuring the clinical viability of the national transition [8,9]. Subsequent viral suppression studies have further confirmed the strong performance of DTG-based regimens and underscored the importance of monitoring regimen transitions using clinical and pharmaceutical indicators [9,10]. These observations reinforce the value of standardised consumption metrics, such as DDD per 1,000 inhabitants per day, to complement programmatic indicators and track shifts in antiretroviral use at scale.

Despite extensive reporting of ART coverage and viral suppression in Tanzania, there has been limited quantification of antiretroviral consumption using the standardised WHO AMC/DDD methodology [8,9]. Applying this approach to ART can reveal regimen utilisation patterns, regional disparities, and the alignment of dispensing data with national treatment coverage. The National AIDS and Sexually Transmitted Infections Control Programme (NASHCoP) in Tanzania has led a nationwide ART scale-up since 2004. It continues to promote rational antiretroviral use through guidelines revisions and service optimisation [11]. However, a national analysis quantifying ART consumption over time and across regions has not yet been conducted.

This study presents the first national-level assessment of antiretroviral consumption in Tanzania, utilising programmatic dispensing data from 2017 to 2021. By expressing utilisation as DDD per 1,000 inhabitants per day, the analysis quantified national and regional trends, captured the transition from efavirenz- to DTG-based therapy, and evaluated geographic variations in ART use. The findings aim to guide evidence-based stewardship, support ART supply planning, and strengthen rational antiretrovirals use [12,13].

## 2. Methodology

### 2.1 Study Design and Setting

This retrospective descriptive study utilised national programmatic data from the NASHCoP to quantify antiretroviral medicine consumption in mainland Tanzania between 2017 and 2021. The analysis included all public and private health facilities that reported dispensing data for antiretroviral medicines through the national HIV Care and Treatment (CTC) and Logistics Management Information Systems. The study followed the World Health Organisation (WHO) methodology for *Antimicrobial Consumption (AMC)*, with results expressed as Defined Daily Doses (DDD) per 1,000 inhabitants per day, the standard international metric for monitoring population-level medicine use [1]. Tanzania is classified as a lower-middle-income country with an adult HIV prevalence of approximately 4.8%, and ART services have been implemented nationwide under the NASHCoP since 2004 [6].

### 2.2 Data Sources

Dispensing data were obtained from the NASHCoP logistics and programmatic databases, which recorded the quantities of antiretroviral medicines dispensed to patients during clinic visits or issued from facility stores. Each record included the ARV drug code or description, pack size and strength, quantity dispensed, the reporting year, and the region of the originating facility. These data represent routine programmatic dispensing and reflect the medicines issued within the national HIV treatment program.

To enable the standardised computation of antiretroviral consumption, the dispensing records were linked with an ARV codes reference table that provided WHO Anatomical Therapeutic Chemical (ATC) classification codes, drug strength expressed in grams per unit, number of units per pack, WHO-defined daily dose (DDD) values, and the therapeutic class of each formulation. A harmonised regimen dataset derived from visit-level dispensing information provided details of the specific formulations and regimen combinations issued during each patient encounter. Annual mid-year population estimates from the National Bureau of Statistics served as denominators for expressing consumption per 1,000 inhabitants per day.

All datasets were linked using common fields, including the drug code and reporting year, and harmonised in Power BI and R. Each dispensed quantity was converted to DDDs by dividing the grams dispensed by the corresponding WHO-DDD value. National and regional ART consumption was expressed as DDDs per 1,000 inhabitants per day, in accordance with the WHO antimicrobial consumption methodology. This harmonisation approach ensured consistent measurements across regions, therapeutic classes, and calendar years.

### 2.3 Data Management and Cleaning

Data cleaning and transformation were performed in Power BI using Power Query (M) to ensure consistency and validity across all the datasets. Records were screened for completeness, and duplicate entries, unmatched drug codes, and dispensing records with invalid quantities or missing key variables were excluded. Regimen identifiers were standardised and mapped to the WHO ATC classifications. Where paediatric formulations or fixed-dose combinations lacked WHO-defined DDD values, proxy DDDs were assigned based on adult equivalents and the national treatment guidelines [1,3]. Population denominators were cross-checked against the official National Bureau of Statistics projections to ensure accurate per-capita consumption calculations. The final analytical dataset, which integrated dispensing volumes, standardised ARV coding, regimen classifications, and annual population estimates, was assessed for internal coherence, outliers, and year-to-year consistency. All transformation steps were scripted and automated to support the reproducibility and auditability of the data processing pipeline.

### 2.4 Computation of Defined Daily Doses (DDD)

Antiretroviral consumption was computed using the WHO antimicrobial consumption methodology. For each dispensing record, the total amount of the active ingredient, in grams, was derived from the quantity dispensed, pack size, and formulation strength. This total amount was then converted into Defined Daily Doses (DDD) by dividing the grams dispensed by the WHO-assigned DDD value for that active ingredient. All calculations were performed in Power BI using Data Analysis Expressions (DAX), which automated the conversion of dispensing volumes into DDDs and linked each record to its corresponding WHO ATC/DDD code. The final indicator, DDDs per 1,000 inhabitants per day, was computed by standardising the total annual DDDs by the mid-year population and dividing by 365 days. This metric reflects the average number of people treated daily per 1,000 population, enabling valid comparisons across years, therapeutic classes and geographic regions. For each dispensing record, the total amount of the active ingredient (in grams) was calculated as follows:

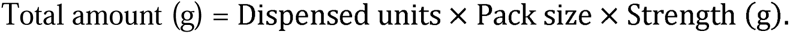

The number of Defined Daily Doses (DDD) was then obtained by dividing the total amount by the WHO-assigned DDD:

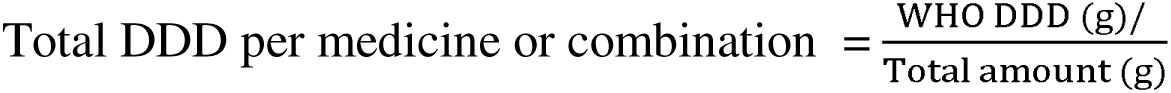

Annual consumption was standardised using mid-year population estimates and expressed as

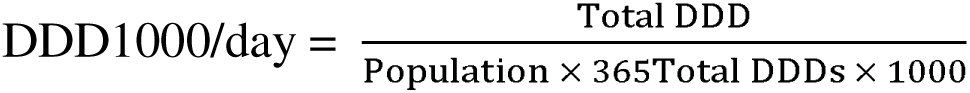

### 2.5 Data Analysis

The Computed DDD values were aggregated by reporting year, therapeutic class, regimen type, and region to describe the national and subnational consumption patterns. Annual and five-year averages were calculated, and a compound annual growth rate (CAGR) was used to summarise the rate of increase in ART utilisation over the study period. Power BI was used for the data transformation and metric computation. Simultaneously, statistical summaries and visualisations—including time-series plots, regional choropleth maps, and ranked bar charts—were generated in R (version 4.x) using the *dplyr*, *sf*, and *ggplot2* packages. This combined analytic workflow ensured both computational accuracy and reproducible visualisation of trends.

### 2.6 Projection to 2027

Short-term projections of national ART consumption were modelled using quadratic regression based on the annual DDD/1,000/day values from 2017 to 2021. Quadratic models were selected because they capture the curvature typically observed in maturing ART programs, allowing for modest acceleration or stabilisation in consumption over time. Projected values were bounded by the upper distribution of observed consumption (95th percentile) to avoid unrealistic extrapolations beyond the expected programmatic saturation level. This modelling approach is consistent with the short-term forecasting methods used in ART demand studies in Sub-Saharan Africa. It provides a reasonable indication of future consumption trajectories under current policies [3,4].

### 2.7 Inclusion Criteria

Records were included if they represented a documented patient visit during which antiretroviral medicines were dispensed and the corresponding quantity was recorded. Visits lacking dispensing activity or missing quantity data were excluded. This ensured that only complete and verifiable dispensing events contributed to the calculation of consumption metrics.

### 2.8 Ethical Considerations

The analysis used aggregated, de-identified programmatic data obtained from the national health information systems and did not involve individual identifiers or direct patient contact. Ethical approval was granted by the Muhimbili University of Health and Allied Sciences Research Ethics Committee (MUHAS-REC), and permission to use the data was obtained from the NASHCoP. All procedures adhered to the national data protection policies and confidentiality standards.

## 3. Results

### 3.1 Overview of ART Service Utilisation, 2017–2021

Between 2017 and 2021, 39,650,725 clinic visits were recorded across all reporting HIV Care and Treatment Clinics, representing 2,108,707 unique patients who received antiretroviral therapy (ART). The annual attendance and utilisation indicators from the national NASHCoP database are summarised in Figure 1 (Panels A–D).

**Figure 1.**
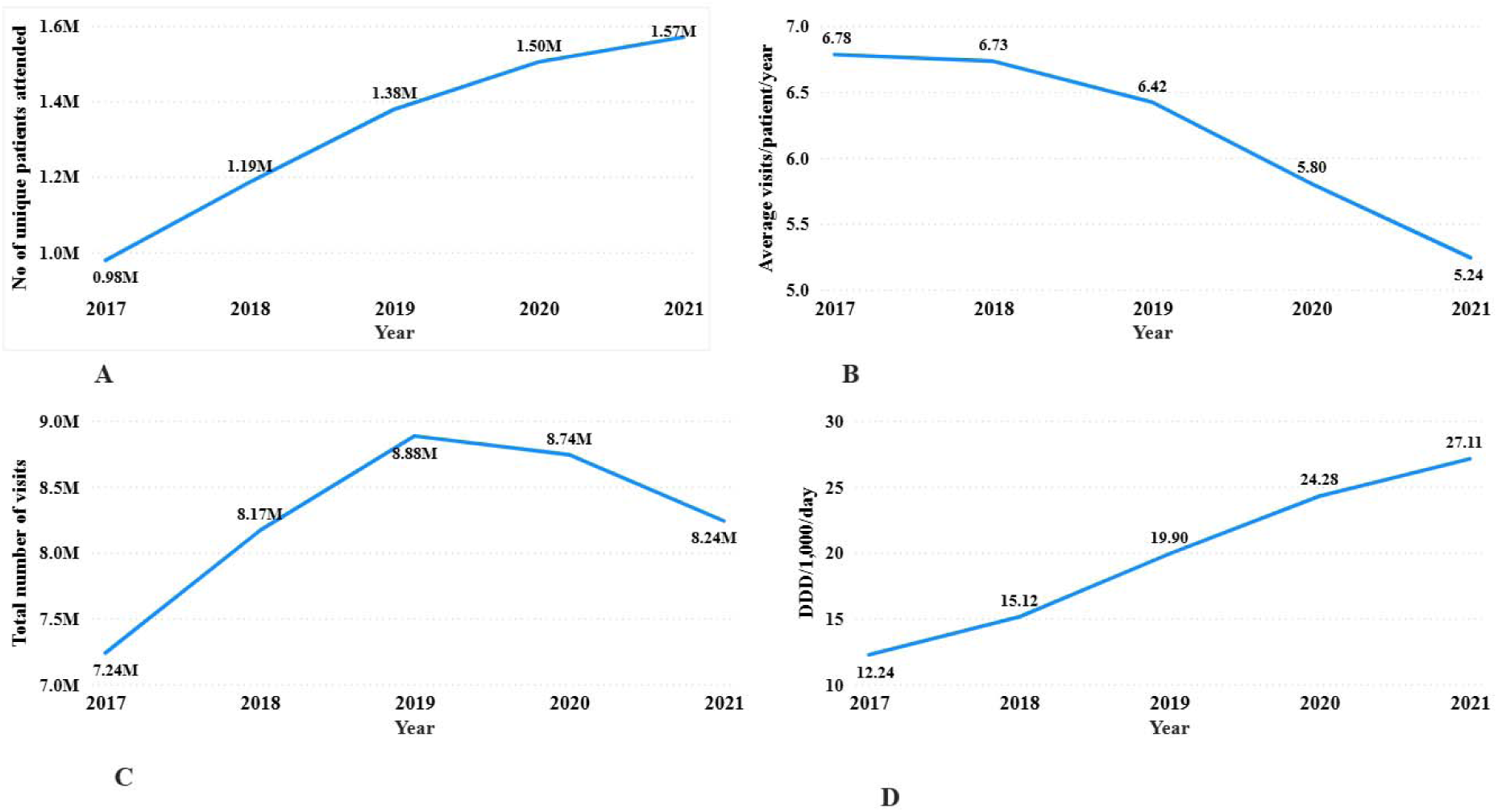
National ART service utilisation and consumption indicators, 2017–2021. Panel A: Number of unique patients visiting CTC facilities. Panel B: Average number of visits per patient per year. Panel C: National ART consumption (DDD per 1,000 inhabitants/day). Panel D: Total number of clinic visits. Data source: National AIDS and Sexually Transmitted Infections Control Programme (NASHCoP), Tanzania Mainland.

The number of unique ART patients increased from 977,725 in 2017 to 1,570,554 in 2021 (Panel A). The average number of visits per patient per year declined from 6.78 in 2017 to 5.24 in 2021 (Panel B). National ART consumption, expressed as Defined Daily Doses (DDD) per 1,000 inhabitants per day, rose from 12.2 DDD/1,000/day in 2017 to 27.1 DDD/1,000/day in 2021, corresponding to a compound annual growth rate of 22.4 % (Panel C). The total number of clinic visits peaked at 8,884,348 in 2019 and declined to 8,238,966 in 2021 (Panel D).

The demographic and dispensing characteristics of the ART records are summarised in Table 1. The distribution of records by gender showed that females accounted for nearly two-thirds of the total dispensing events. Most visits were routine treatment encounters (visit types 2–4), and more than 99 % of all entries corresponded to first-line regimens.

**Table 1:**
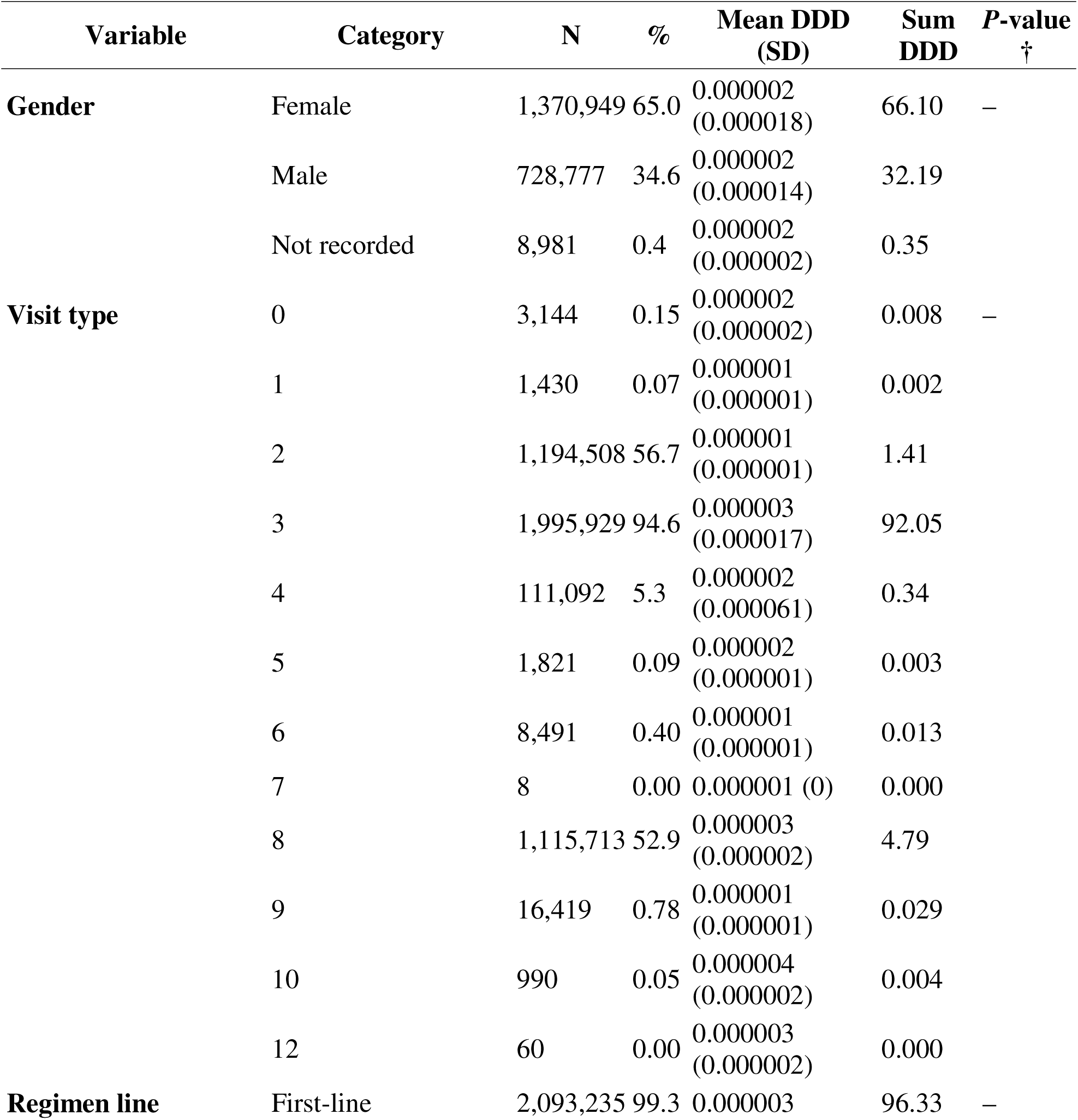
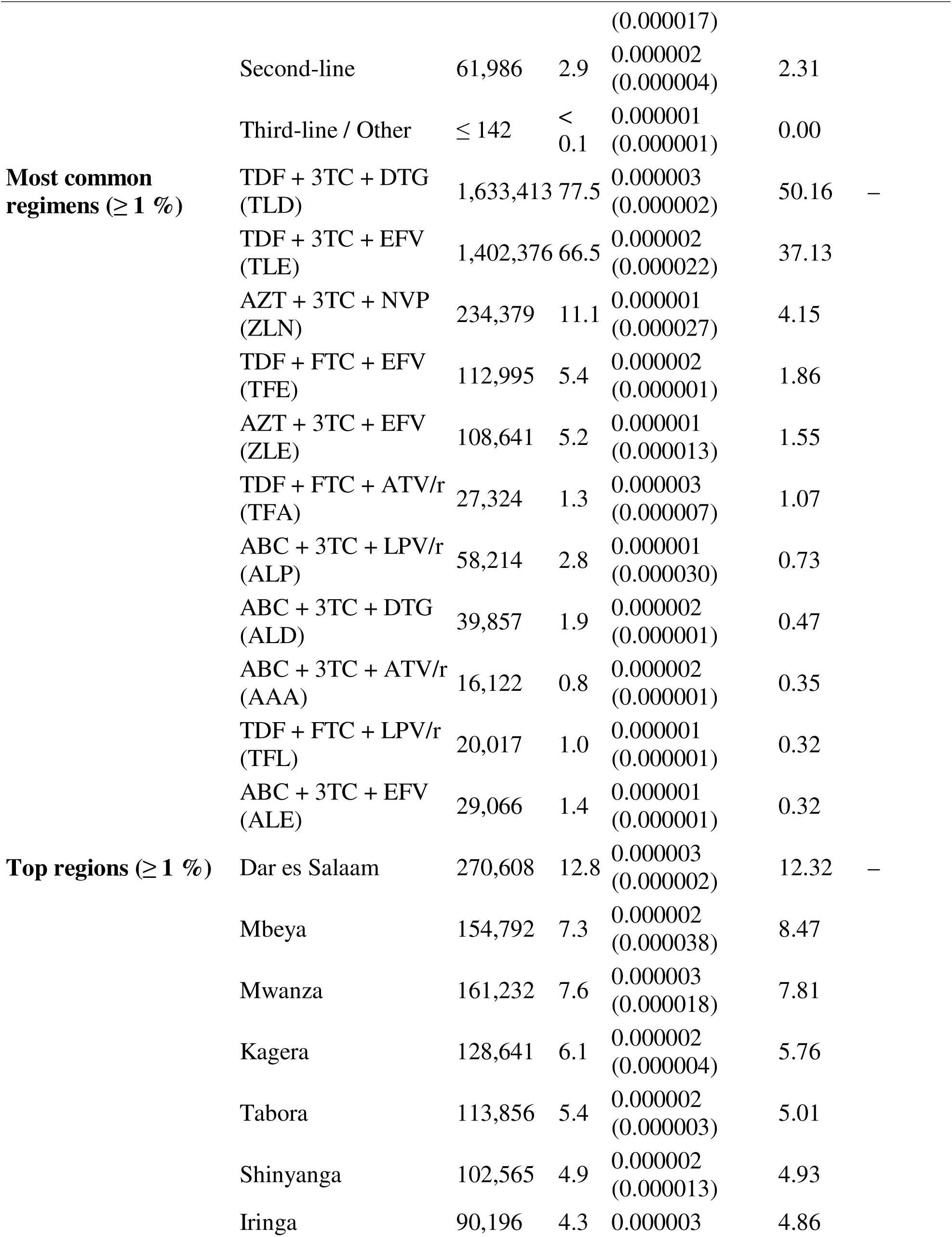
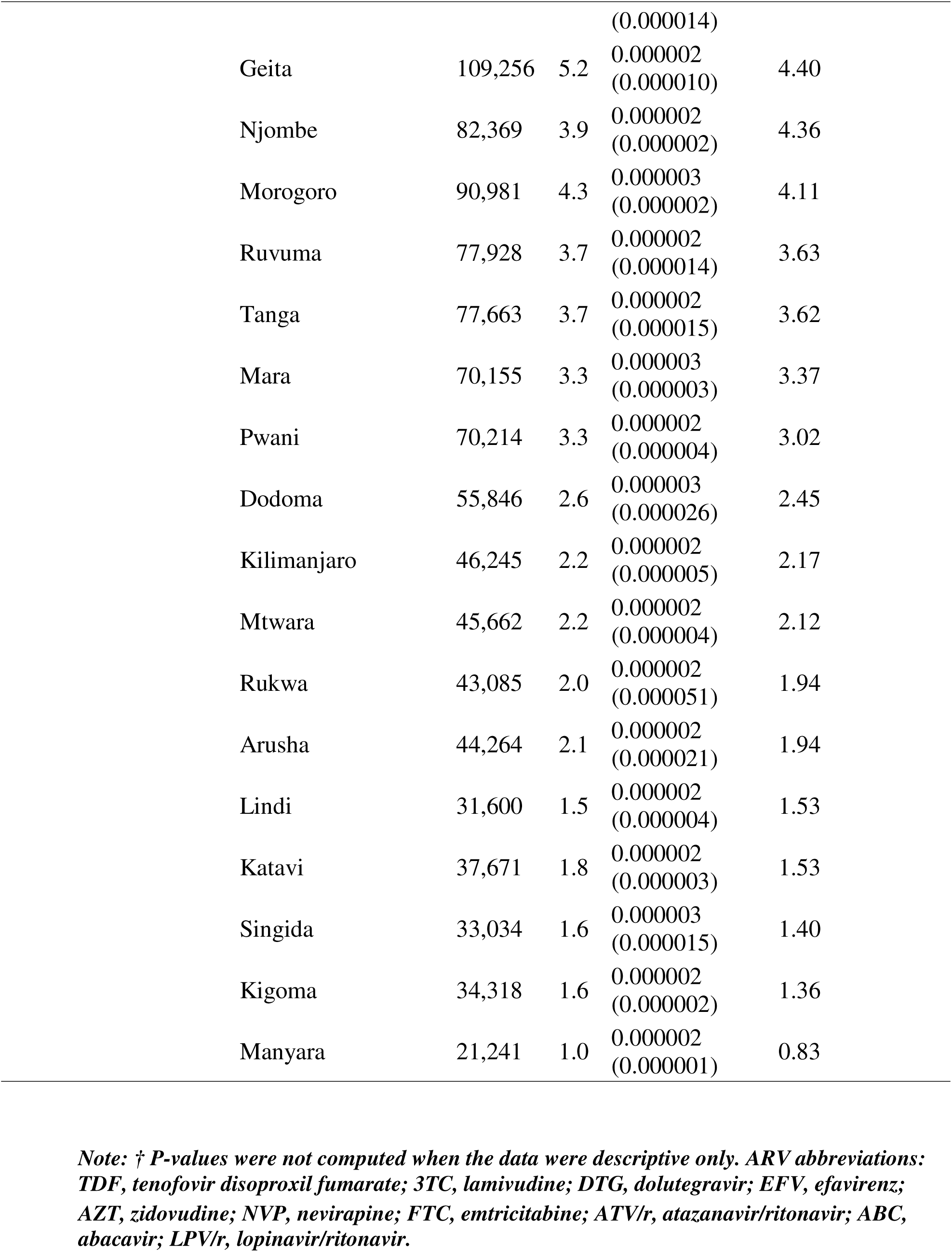
Demographic, visit, regimen line, and regional characteristics of ART dispensing records, Tanzania 2017–2021.

DTG-based combinations (TDF + 3TC + DTG) represented the largest proportion of dispensed ART regimens, followed by efavirenz-based (TDF + 3TC + EFV) and zidovudine-based (AZT + 3TC + NVP) regimens. Some regimens were less common (Supplementary Table S1). The regional representation was broad, with the highest dispensing volumes recorded in Dar es Salaam, Mbeya, and Mwanza.

Subsequent analyses disaggregated these data to show regional variations in ART consumption (Figure 2) and temporal trends by regimen class, namely, non-nucleoside reverse transcriptase inhibitors (NNRTIs), protease inhibitors (PIs), and integrase strand transfer inhibitors (INSTIs) (Figure 3).

**Figure 2.**
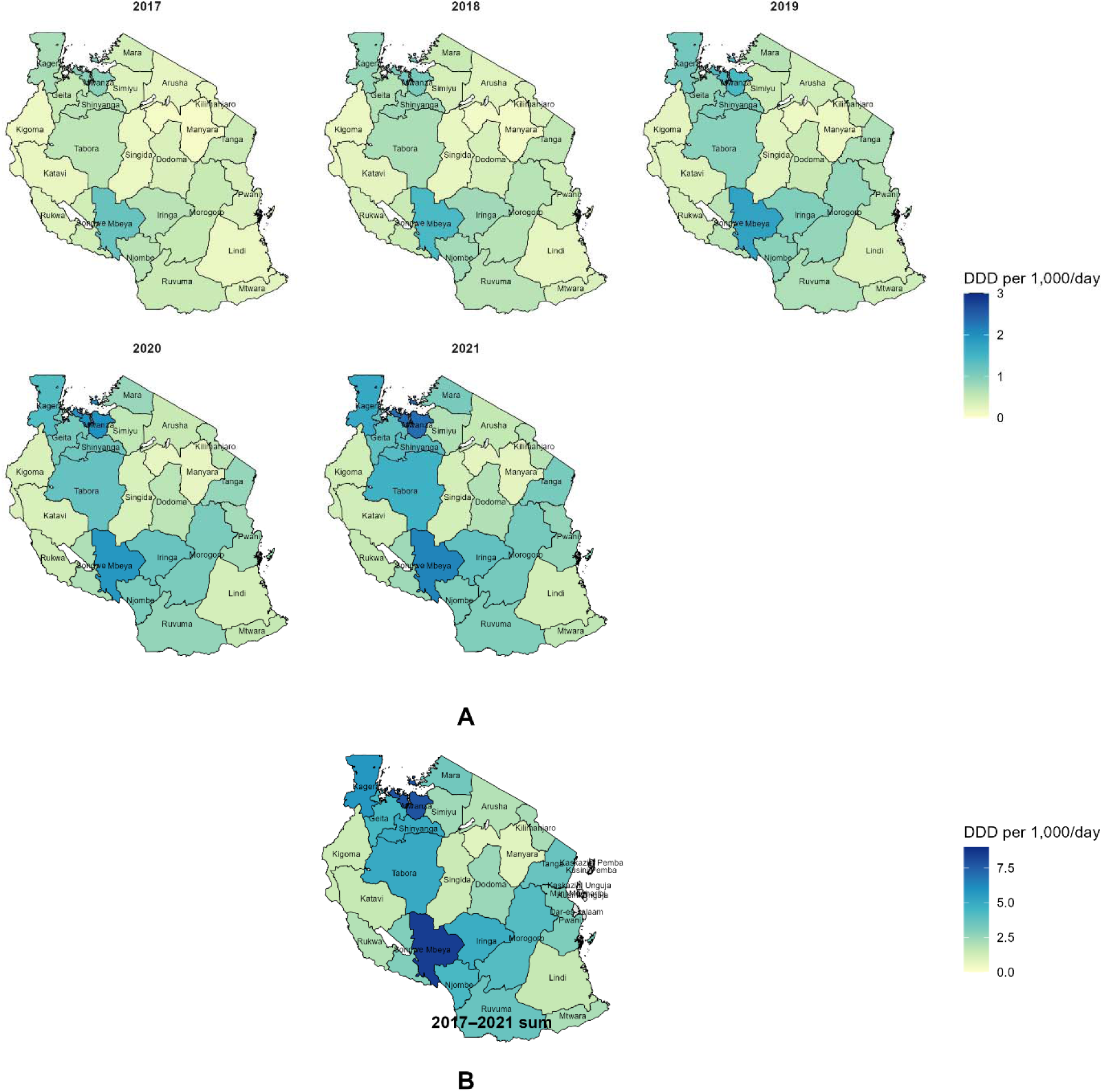
Regional antiretroviral therapy (ART) consumption in Tanzania, 2017–2021. Panel A: Annual maps of ART consumption expressed as defined daily doses (DDD) per 1,000 inhabitants per day, showing progressive expansion of therapy coverage and Panel B: Cumulative consumption across the five years (2017–2021 sum). Data source: National AIDS and Sexually Transmitted Infections Control Programme (NASHCoP), Tanzania Mainland.

**Figure 3.**
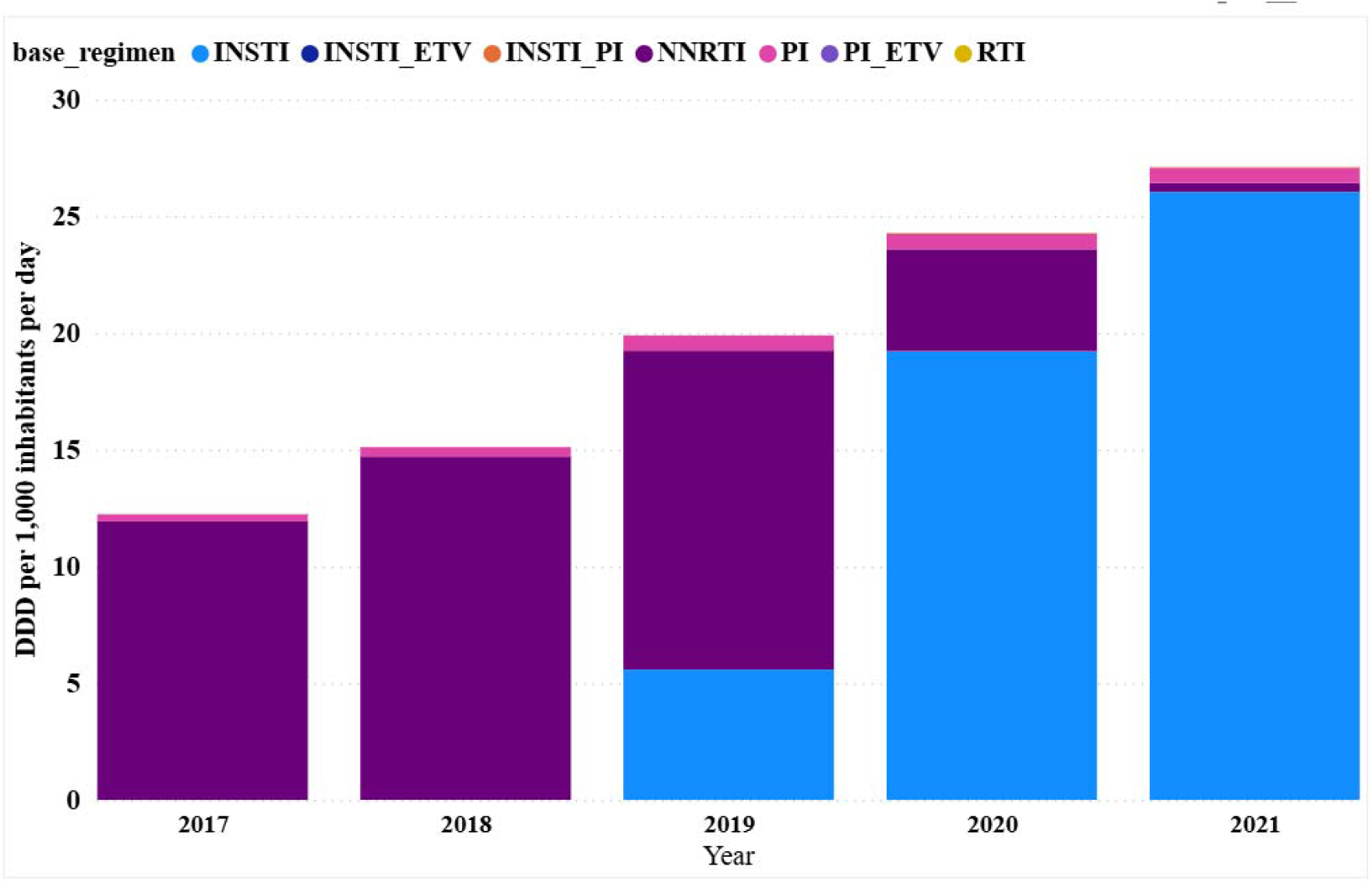
Annual national ART consumption by regimen class in Tanzania, 2017–2021. The lines represent the defined daily doses (DDD) per 1,000 inhabitants per day for non-nucleoside reverse transcriptase inhibitors (NNRTIs), integrase strand transfer inhibitors (INSTIs), and protease inhibitors (PIs). Data source: National AIDS and Sexually Transmitted Infections Control Programme (NASHCoP), Tanzania Mainland.

### 3.5 Regional Variation in ART Consumption

Regional-level analysis (Figure 2) showed moderate spatial variation in antiretroviral therapy (ART) consumption across mainland Tanzania. Urban regions such as Dar es Salaam, Mwanza, and Arusha recorded the highest defined daily doses (DDD) per 1,000 inhabitants per day, whereas Rukwa, Katavi, and Lindi reported the lowest levels. The cumulative five-year map (Figure 2B) confirms this pattern, with higher overall consumption concentrated in urban and lake-zone regions. When normalised to the population, the differences between regions narrowed, indicating broadly comparable access to ART nationwide. Additional numerical values corresponding to the regional maps are provided in Supplementary Table S2, which lists the annual and cumulative ART consumption (DDD per 1,000 inhabitants per day) for all 26 regions of the Tanzanian Mainland from 2017 to 2021.

### 3.6 Temporal Trends in ART Regimen Classes

National trends in regimen class utilisation (Figure 3) indicate a marked shift from non-nucleoside reverse transcriptase inhibitor (NNRTI)-based regimens to integrase strand transfer inhibitor (INSTI)-based therapy between 2017 and 2021. NNRTI use declined from 11.9 to 0.4 DDD per 1,000 inhabitants/day, whereas INSTI-based consumption increased more than tenfold, from 0.002 to 26.0 DDD per 1,000 inhabitants/day. Protease inhibitor (PI) use remained stable at approximately 0.3–0.7 DDD per 1,000 inhabitants per day throughout the study period, and other regimen categories (RTI, INSTI_PI, INSTI_ETV, PI_ETV) contributed minimally. These results reflect the national transition to DTG-based first-line therapy, which was initiated in 2019.

Together, these temporal and spatial analyses highlight the rapid national transition to DTG-based regimens and regional heterogeneity in ART distribution patterns across Tanzania. The subsequent discussion interprets these findings in relation to shifts in the national treatment policy and evolving program coverage during the study period.

### 3.7 National Forecast of ART Consumption

Forecast modelling using national annual ART consumption data (2017–2021) predicted a continued upward trajectory until 2025 (Figure 4). The fitted polynomial trend indicated a steady increase in defined daily doses (DDD) per 1,000 inhabitants per day, consistent with the sustained expansion of DTG-based therapy and the national program scale-up. The projected 2025 value approached the upper range of the 95% prediction interval, suggesting that ART coverage in Tanzania is likely to continue growing under the current policy implementation.

**Figure 4.**
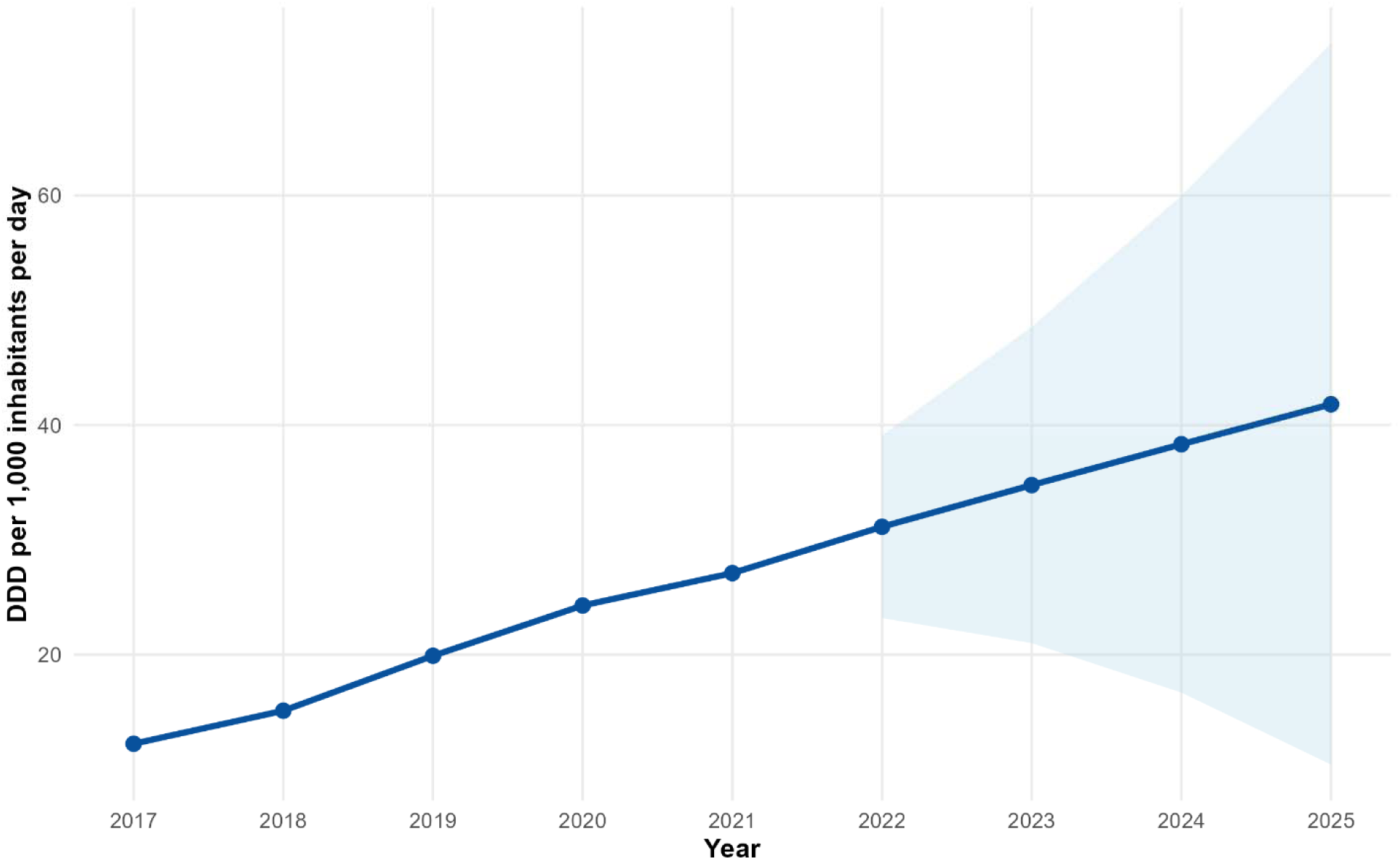
Observed (2017–2021) and projected (2022–2025) national antiretroviral therapy (ART) consumption in Tanzania. The solid line represents the fitted values from the second-order polynomial model, whereas the shaded area indicates the 95% prediction interval. Values are expressed as defined daily doses (DDD) per 1,000 inhabitants/day. Data source: National AIDS and Sexually Transmitted Infections Control Programme (NASHCoP), Tanzania Mainland.

### 3.8 Summary of Findings

Between 2017 and 2021, national antiretroviral therapy (ART) consumption in Tanzania increased from 12.2 to 27.1 defined daily doses (DDD) per 1,000 inhabitants per day, with a five-year mean of 19.7 DDD per 1,000 inhabitants/day. Nucleoside/nucleotide reverse transcriptase inhibitors (NRTIs) remained the largest therapeutic class, whereas DTG-based combinations have become the predominant regimens since 2019. Regional analyses showed consistent upward trends across all zones, with higher consumption in the urban and lake zone regions. Forecast modelling suggested continued moderate national growth through 2025, reflecting the sustained expansion of ART access and the integration of newer regimen classes.

## 4. Discussion

This study presents the first national analysis of antiretroviral therapy (ART) consumption in Tanzania using the WHO Defined Daily Dose (DDD) methodology. It offers a standardised measure of ART utilisation that complements routine HIV program metrics. By leveraging programmatic dispensing data from 2017 to 2021 and applying a validated AMC framework, the analysis captured both temporal and regional patterns during a period of substantial treatment optimisation, including the national transition to DTG-based regimens. These findings provide quantitative insights into the scale, distribution, and trajectory of ART use in Tanzania, offering a foundation for strengthening data-driven planning, stewardship, and evaluation within the national HIV response.

### 4.1 Main Findings

This national assessment revealed a sustained increase in ART consumption in Tanzania between 2017 and 2021, rising from 12.2 to 27.1 DDD per 1,000 inhabitants per day, with a five-year mean of 19.7 DDD per 1,000 inhabitants per day. Nucleoside/nucleotide reverse transcriptase inhibitors (NRTIs) have been the therapeutic backbone for many years. In contrast, integrase strand transfer inhibitor (INSTI)-based combinations have expanded rapidly since 2019, following the national implementation of DTG as first-line therapy in line with WHO guidelines [3,9,11,14]. Although regional differences in consumption persisted, particularly between urban/lake-zone regions and several southern districts, the overall variation narrowed over time, suggesting progressively harmonised access to ART. Forecast modelling suggests that national consumption is likely to continue rising moderately through 2025, reflecting ongoing program expansion and demographic growth.

Service utilisation indicators support this pattern, with rising numbers of unique ART patients from 2017 to 2021, consistent with the nationwide scale-up under the “Treat All” strategy. Concurrently, the decline in average clinic visits per patient per year occurred during the period of programmatic efforts to streamline service delivery, including the adoption of multi-month dispensing and differentiated service delivery models (DSDMs). The rise in total clinic visits between 2017 and 2019, followed by a decline in 2020–2021, occurred alongside regimen optimisation activities and broader service reorganisation, potentially influenced by program efficiencies and COVID-19–related shifts in patient flow.

### 4.2 Comparison with Previous Studies

The upward trajectory in ART consumption observed in this study is consistent with earlier national evidence showing the progressive scale-up of HIV treatment in Tanzania and the feasibility of applying WHO DDD methods to routine program data to track temporal changes in medicine use [7]. The rapid rise in integrase inhibitor consumption aligns with clinical experience from the early DTG rollout in Dar es Salaam, where high adherence and favourable outcomes were reported following regimen transition [10], and with more recent observations of high viral suppression and emerging DTG-associated mutations in treatment-experienced adults [8]. Together, these findings provide a clinical context for the substantial increase in INSTI-based DDDs documented in this study.

At the regional level, the observed patterns resemble trends reported across sub-Saharan Africa, where national transitions to DTG in Kenya, Uganda and South Africa have resulted in rising total DDDs and greater regimen standardisation [1,5,11,12,14].

Broader global analyses similarly show continued expansion of ART coverage and evolving treatment compositions as programs align with updated WHO guidance [1] (Lancet HIV 2024). International antimicrobial consumption research supports these patterns; for example, Kazakhstan reported marked increases in antiviral use following the introduction of newer drug classes [6]. Earlier global forecasting anticipated these shifts, projecting increases in antiretroviral demand and a transition away from older regimens, such as stavudine, toward more effective combinations [5,13,15]. More recent modelling has also highlighted the influence of treatment guidelines on ART initiation dynamics and national supply needs [5,16,17].

Taken together, these studies reflect a consistent global narrative: treatment optimisation policies, such as Tanzania’s transition to DTG-based regimens, produce measurable changes in national antiretroviral consumption profiles. The present analysis situates Tanzania within this broader trajectory and underscores the importance of ongoing consumption monitoring to support the forecasting, procurement and evaluation of guideline implementation.

### 4.3 Regional Variation and Access

Although variations in ART consumption remained across regions, the overall disparity decreased over time. Higher DDD/1,000/day values in Dar es Salaam, Mwanza, Arusha, and other urban or lake-zone regions reflect areas with larger patient populations and more established treatment infrastructures. Lower consumption in several western and southern regions may partly reflect facility density and reporting completeness rather than restricted access to care. Future analyses linking DDD data to viral suppression, retention in care, or geographic service coverage indicators will help clarify whether the remaining differences reflect operational challenges or underlying epidemiological patterns of HIV.

### 4.4 Programmatic and Policy Implications

This study demonstrates that the DDD/1 000/day metric can be integrated into routine national monitoring as a complementary indicator of ART utilisation and regimen uptake. Routine computation of AMC indicators would enable NASHCoP to monitor trends in near real-time, validate programmatic ART coverage reports, and improve procurement quantification and forecasting. The rapid shift toward DTG-based therapy confirms the effective implementation of the 2019–2021 optimisation roadmap and alignment with WHO recommendations. Linking AMC measures with viral load suppression, early warning indicators, and HIV drug resistance surveillance could strengthen pharmacovigilance, support rational ARV use, and improve early detection of supply imbalances.

### 4.5 Strengths of the Study

This is the first nationwide application of the WHO DDD methodology to antiretroviral medicines in Tanzania. The analysis was based on routine dispensing data, capturing real-world utilisation rather than procurement volumes, and used standardised population denominators for national and regional comparisons. The reproducible analytic framework, including Power BI computation and GIS-based regional mapping, provides a practical model for integrating AMC metrics into ongoing HIV program monitoring. Including a short-term projection provides insights into future program stability and resource requirements.

### 4.6 The Future Directions

Integrating AMC indicators with viral load suppression, retention data, and HIV drug resistance surveillance would enhance the early detection of regimen performance issues and strengthen forecasts. Embedding AMC dashboards within NASHCoP’s electronic logistics platforms would facilitate routine monitoring at the national and regional scale. Expanding surveillance to private sector providers, military services, and donor-supported supply chains would enable a more comprehensive national estimate of ART consumption in the country.

One limitation of this analysis is the use of proxy DDD values for certain paediatric formulations and fixed-dose combinations, which may slightly reduce the comparability across products. Additionally, population estimates were based on mid-year projections, which may have introduced minor uncertainty in the per-capita calculations. Finally, we did not associate consumption with adherence, wastage, viral load suppression, or drug resistance outcomes.

### 4.8 Conclusions

Antiretroviral consumption in Tanzania increased steadily from 2017 to 2021, driven by the continued expansion of ART services and rapid transition to DTG-based regimens. Applying the WHO DDD methodology to programmatic dispensing data provides a standardised, feasible approach for monitoring ART use and evaluating regimen transitions. These findings support the further integration of AMC indicators into national HIV program evaluation frameworks to promote efficient supply planning and the rational use of antiretrovirals.

## 5. Conclusion and Recommendations

### 5.1 Conclusion

This study provides the first national quantification of antiretroviral medicine consumption in Tanzania using the WHO’s Defined Daily Dose (DDD) per 1,000 inhabitants per day metric. Analysis of NASHCoP dispensing data from 2017 to 2021 showed a steady rise in ART utilisation, consistent with the expansion of treatment coverage under the national “Treat All” strategy. The transition from efavirenz-based to DTG-based combinations represents a major shift in treatment patterns and reflects strong national adherence to the WHO guidelines. Regional variations in consumption were modest and generally aligned with known differences in HIV burden and service availability. Overall, the DDD/1 000/day indicator provided a practical and cost-effective metric for monitoring ART utilisation.

### 5.2 Recommendations

The routine integration of the DDD per 1,000 inhabitants per day metric into NASHCoP’s monitoring framework would strengthen the national tracking of ART utilisation and regimen changes. Linking AMC indicators with viral load suppression and HIV drug resistance surveillance would enhance the early detection of treatment performance issues and improve forecasts. Strengthening data completeness and harmonisation within the eLMIS and CTC systems, and expanding analyses to include private and non-governmental providers, would produce more comprehensive national estimates. Finally, building technical capacity in AMC computation and data visualisation among regional and national program staff would support sustained evidence-based stewardship of antiretroviral medicines.

## Acknowledgements

The authors acknowledge the National AIDS and Sexually Transmitted Infections Control Programme (NASHCoP), Ministry of Health, Tanzania, for providing access to the national ART dispensing data. We thank the regional and facility-level teams involved in the routine reporting of HIV care and treatment. We also extend our appreciation to the Muhimbili University of Health and Allied Sciences (MUHAS) Directorate of Research and Publications for their institutional support throughout this analysis.

## Ethical Approval

This study used secondary, de-identified, and aggregated program data. Ethical clearance was granted by the Muhimbili University of Health and Allied Sciences Research Ethics Committee (MUHAS-REC), and NASHCoP provided written permission for the use of data. No individual-level identifiers were accessed, and all analyses followed the national data protection policies and established ethical standards.

## Data Availability Statement

All data produced in the present study are available upon reasonable request to the authors.

## Supplementary Tables

**Supplementary Table S1:** Regimens accounting for < 1 % of total ART dispensing records, Tanzania 2017–2021.

| Regimen | N | % | Mean DDD (SD) | Sum DDD |
| --- | --- | --- | --- | --- |
| ABC + 3TC + ATV/r | 16,122 | 0.8 | 0.000002 (0.000001) | 0.35 |
| TDF + FTC + LPV/r | 20,017 | 1.0 | 0.000001 (0.000001) | 0.32 |
| ABC + 3TC + EFV | 29,066 | 1.4 | 0.000001 (0.000001) | 0.32 |
| AZT + 3TC + LPV/r | 7,188 | 0.3 | 0.000002 (0.000001) | 0.13 |
| AZT + 3TC + DTG | 13,089 | 0.6 | 0.000003 (0.000002) | 0.13 |
| AZT + 3TC + ATV/r | 6,197 | 0.3 | 0.000001 (0.000001) | 0.09 |
| TDF + FTC + DTG | 10,725 | 0.5 | 0.000003 (0.000002) | 0.07 |
| Other second-line | 5,905 | 0.3 | 0.000002 (0.000001) | 0.07 |
| DTG + ABC/3TC + ATV/r | 1,271 | 0.1 | 0.000002 (0.000001) | 0.02 |
| TDF + FTC + NVP | 1,206 | 0.1 | 0.000002 (0.000001) | 0.01 |
| ABC + 3TC + NVP | 520 | 0.02 | 0.000002 (0.000001) | 0.00 |
| TDF + 3TC + ATV/r | 220 | 0.01 | 0.000002 (0.000001) | 0.00 |
| TDF + 3TC + LPV/r | 320 | 0.02 | 0.000001 (0.000001) | 0.00 |
| TDF + 3TC + NVP | 433 | 0.02 | 0.000002 (0.000001) | 0.00 |
| d4T + 3TC + NVP | 395 | 0.02 | 0.000001 (0.000001) | 0.00 |
| ABC + DDL + LPV/r | 94 | 0.00 | 0.000001 (0.000001) | 0.00 |
| ABC + DDL + SQV/r | 120 | 0.01 | 0.000001 (0.000001) | 0.00 |
| d4T + 3TC + EFV | 56 | 0.00 | 0.000001 (0.000001) | 0.00 |
| ABC + DDL + NFV | 74 | 0.00 | 0.000002 (0.000001) | 0.00 |
| DTG + ATV/r + ETV | 15 | 0.00 | 0.000002 (0.000001) | 0.00 |
| DTG + DRV/r + ETV | 19 | 0.00 | 0.000001 (0) | 0.00 |
| Other third-line | 47 | 0.00 | 0.000002 (0.000001) | 0.00 |
| RAL + DRV/r + ETV | 3 | 0.00 | 0.000001 (0) | 0.00 |
| AZT + 3TC prophylaxis | 4 | 0.00 | 0.000001 (0.000001) | 0.00 |
| AZT prophylaxis | 3 | 0.00 | 0.000001 (0.000001) | 0.00 |
| d4T + 3TC + LPV/r | 4 | 0.00 | 0.000001 (0.000001) | 0.00 |
| RAL + LPV/r + TDF/FTC | 1 | 0.00 | 0.000001 (0) | 0.00 |
| AZT + DDL + NVP | 1 | 0.00 | 0.000001 (0) | 0.00 |

**Abbreviations:** *TDF*, tenofovir disoproxil fumarate; *3TC*, lamivudine; *DTG*, dolutegravir; *FV*, efavirenz; *AZT*, zidovudine; *NVP*, nevirapine; *FTC*, emtricitabine; *ATV/r*, atazanavir/ritonavir; *LPV/r*, lopinavir/ritonavir; *ABC*, abacavir; *d4T*, stavudine; *DDL* (*ddI*),
*didanosine; SQV/r, saquinavir/ritonavir; NFV, nelfinavir; DRV/r, darunavir/ritonavir; ETV, etravirine; RAL, raltegravir.*

**Supplementary Table S2:** Regional annual and cumulative ART consumption (DDD per 1,000 inhabitants per day), 2017-2021.

| **Region** | **2017** | **2018** | **2019** | **2020** | **2021** | **Five-year cumulative** |
| --- | --- | --- | --- | --- | --- | --- |
| Dar es Salaam | 8.5 | 10.9 | 13.1 | 15.0 | 17.4 | **64.9** |
| Mbeya | 6.2 | 7.8 | 9.2 | 10.2 | 11.7 | **45.1** |
| Mwanza | 5.7 | 7.1 | 8.5 | 9.8 | 11.0 | **42.1** |
| Kagera | 4.1 | 5.2 | 6.4 | 7.3 | 8.4 | **31.4** |
| Tabora | 3.7 | 4.6 | 5.7 | 6.5 | 7.3 | **27.8** |
| Shinyanga | 3.6 | 4.5 | 5.5 | 6.2 | 7.1 | **26.9** |
| Iringa | 3.4 | 4.3 | 5.4 | 6.0 | 6.9 | **26.0** |
| Geita | 3.3 | 4.1 | 5.1 | 5.8 | 6.7 | **24.9** |
| Njombe | 3.2 | 4.0 | 4.9 | 5.5 | 6.3 | **24.0** |
| Morogoro | 3.1 | 3.9 | 4.9 | 5.6 | 6.4 | **23.9** |
| Ruvuma | 2.9 | 3.6 | 4.6 | 5.3 | 6.1 | **22.5** |
| Tanga | 2.9 | 3.6 | 4.6 | 5.2 | 5.9 | **22.2** |
| Mara | 2.7 | 3.4 | 4.3 | 5.0 | 5.7 | **21.1** |
| Pwani | 2.6 | 3.3 | 4.1 | 4.8 | 5.5 | **20.3** |
| Dodoma | 2.1 | 2.8 | 3.6 | 4.3 | 5.0 | **17.8** |
| Kilimanjaro | 1.9 | 2.6 | 3.4 | 4.0 | 4.6 | **16.5** |
| Mtwara | 1.9 | 2.5 | 3.2 | 3.8 | 4.5 | **15.9** |
| Rukwa | 1.7 | 2.3 | 3.0 | 3.6 | 4.3 | **14.9** |
| Arusha | 1.7 | 2.3 | 2.9 | 3.5 | 4.2 | **14.6** |
| Lindi | 1.3 | 1.8 | 2.4 | 2.9 | 3.5 | **11.9** |
| Katavi | 1.3 | 1.8 | 2.3 | 2.8 | 3.3 | **11.5** |
| Singida | 1.2 | 1.7 | 2.3 | 2.8 | 3.3 | **11.3** |
| Kigoma | 1.2 | 1.6 | 2.1 | 2.6 | 3.0 | **10.5** |
| Manyara | 0.7 | 0.9 | 1.2 | 1.5 | 1.8 | **6.1** |
| Songwe | 2.5 | 3.2 | 4.0 | 4.7 | 5.4 | **19.8** |
| Simiyu | 2.4 | 3.1 | 3.8 | 4.5 | 5.2 | **19.0** |
| **National mean** | **12.2** | **15.1** | **19.9** | **24.3** | **27.1** | **98.6** |

*DDD: Defined Daily Dose; values represent DDD per 1,000 inhabitants per day.*
*Data source: National AIDS and Sexually Transmitted Infections Control Programme (NASHCoP), Tanzania Mainland.*

